# Genome-wide association studies and multi-omics integrative analysis reveal novel loci and their molecular mechanisms for circulating polyunsaturated, monounsaturated, and saturated fatty acids

**DOI:** 10.1101/2024.11.11.24317110

**Authors:** Yitang Sun, Huifang Xu, Kaixiong Ye

## Abstract

Previous genome-wide association studies (GWAS) have identified genetic loci associated with the circulating levels of FAs, but the biological mechanisms of these genetic associations remain largely unexplored. Here, we conducted GWAS to identify additional genetic loci for 19 circulating fatty acid (FA) traits in UK Biobank participants of European ancestry (N = 239,268) and five other ancestries (N = 508 – 4,663). We leveraged the GWAS findings to characterize genetic correlations and colocalized regions among FAs, explore sex differences, examine FA loci influenced by lipoprotein metabolism, and apply statistical fine-mapping to pinpoint putative causal variants. We integrated GWAS signals with multi-omics quantitative trait loci (QTL) to reveal intermediate molecular phenotypes mediating the associations between the genetic loci and FA levels. Altogether, we identified 215 significant loci for polyunsaturated fatty acids (PUFAs)-related traits in European participants, 163 loci for monounsaturated fatty acids (MUFAs)-related traits, and 119 loci for saturated fatty acids (SFAs)-related traits, including 70, 61, and 54 novel loci, respectively. A novel locus for total FAs, the percentage of omega-6 PUFAs in total FAs, and total MUFAs (around genes *GSTT1/2/2B*) overlapped with QTL signals for all six molecular phenotypes examined, including gene expression, protein abundance, DNA methylation, splicing, histone modification, and chromatin accessibility. Across 19 FA traits, 65% of GWAS loci overlapped with QTL signals for at least one molecular phenotype. Our study identifies novel genetic loci for circulating FA levels and systematically uncovers their underlying molecular mechanisms.

## Introduction

Polyunsaturated fatty acids (PUFAs), monounsaturated fatty acids (MUFAs), and saturated fatty acids (SFAs) have been implicated in various diseases.^1–3^ Evidence from epidemiological and genetic studies indicates that PUFAs, especially omega-3 PUFAs, are associated with reduced risks of various diseases or mortality by modifying metabolism and inflammation.^4–7^ Circulating fatty acid (FA) levels are influenced by environmental and genetic factors and characterized by sex- and ancestry-specific patterns.^8–11^ Dietary intake, socioeconomic status, physical inactivity, cigarette smoking, and alcohol consumption are common environmental factors that influence FA levels.^12^ Although hundreds of genetic loci associated with FAs have been identified by genome-wide association studies (GWAS),^13–38^ the biological mechanisms of these genetic associations remain largely unexplored. Moreover, these genetic loci were mainly discovered in the European (EUR) population and were focused on autosomes. Extending GWAS to non-EUR populations and sex chromosomes will offer a comprehensive understanding of the genetic architecture of circulating FAs.

In this study, we perform the GWAS for 19 FA traits related to PUFAs, MUFAs, and SFAs to identify more novel genetic loci. We conducted overall and sex-specific GWAS using 250,101 individuals of EUR ancestry and five other ancestries from the UK Biobank (UKB). Leveraging the GWAS summary statistics, we characterized heritability for these FA traits and examined their shared genetic basis with genetic correlation and colocalization analysis. Candidate causal variants were pinpointed with statistical fine-mapping. Finally, we integrated GWAS signals with multi-omics data to identify the mediating molecular phenotypes and provide mechanistic insights into the molecular mechanisms underlying the GWAS signals.

## Methods

### Participants

The UKB cohort is a prospective population-based study of ∼500,000 participants from across the United Kingdom, aged between 37 and 73 years at recruitment from 2006 to 2010.^39^ Of the participants with both phenotype and genotype data, we excluded those who have withdrawn consent, mismatched information between self-reported and genetic sex, sex chromosome aneuploidy, or are outliers for heterogeneity and missing genotype rate. Genetic ancestry groups have been previously defined in the Pan-ancestry genetic analysis of the UK Biobank (Pan-UKB).^40^ Together, we included up to 239,268 participants of EUR ancestry in this study. We also included participants of African (AFR) (N = 3,352), admixed American (AMR) (N = 508), Central/South Asian (CSA) (N = 4,663), East Asian (EAS) (N = 1,445), and Middle Eastern (MID) (N = 865) ancestries. Table S1 provides the overall and sex-stratified characteristics of participants in each ancestry group. This study received ethical approval from the National Health Service North West Centre for Research Ethics Committee, and all participants provided informed consent. Data from the UKB resource were accessed under application number 48818.

### Phenotypes – circulating FAs

Plasma FAs were measured in a randomly selected subset of 274,020 participants by Nightingale Health using nuclear magnetic resonance (NMR) spectroscopy.^41^ All FA levels and their ratios were normalized within each ancestry group using the rank-based inverse normal transformation in overall GWAS.^42^ This inverse normal transformation was applied separately for males and females to account for sex-specific differences in FA distributions. In total, 20 metabolic measurements were initially analyzed in the GWAS, including total FAs, 5 PUFA absolute concentrations (i.e., total PUFAs, omega-3 PUFAs, omega-6 PUFAs, docosahexaenoic acid (DHA), and linoleic acid (LA)), their relative percentages in total FAs (i.e., PUFAs%, omega-3%, omega-6%, DHA%, and LA%), the ratio of omega-3 to omega-6 PUFAs (omega- 3/omega-6), the ratio of omega-6 to omega-3 PUFAs (omega-6/omega-3), monounsaturated fatty acids (MUFAs), saturated fatty acids (SFAs), MUFAs%, SFAs%, PUFAs/MUFAs, PUFAs/SFAs, and degree of unsaturation. Both the omega-3/omega-6 ratio and omega-6/omega- 3 ratio were included in our GWAS due to different preferences in the field, but they revealed the same genetic loci in opposite effect directions. In additional analyses, we focused on omega- 3/omega-6 and did not include omega-6/omega-3, resulting in a total of 19 plasma metabolic measurements. FA traits were categorized into PUFA-related traits (total PUFAs, omega-3 PUFAs, omega-6 PUFAs, DHA, LA, PUFAs%, omega-3%, omega-6%, DHA%, LA%, omega- 3/omega-6, PUFAs/MUFAs, PUFAs/SFAs, and degree of unsaturation), MUFA-related traits (total MUFAs, MUFAs%, and PUFAs/MUFAs), and SFA-related traits (total SFAs, SFAs%, and PUFAs/SFAs).

### Genotyping and quality control

Full details about genotyping, imputation, and genotype-based quality control (QC) have been described elsewhere.^43^ Briefly, we excluded variants with imputation quality score < 0.3, minor allele frequency < 0.001, genotype missingness per variant > 0.05, and Hardy-Weinberg test *P*-value < 1 × 10^−8^. We also confirmed that no individuals have genotype missingness > 0.05. For the non-pseudoautosomal region of the X chromosome (GRCh37; chrX:2699520- 154931044), males were treated as homozygotes of the reference or effect allele, and coded as 0 or 2, respectively.

### Genome-wide association studies (GWAS)

Overall GWAS were conducted for each ancestry group and followed by sex-stratified GWAS. We carried out GWAS in participants of EUR ancestry using fastGWA from the GCTA toolbox (v.1.94.1), which controls for familial relatedness using a sparse genetic relationship matrix (GRM) with a default threshold of 0.05 based on slightly linkage disequilibrium (LD)- pruned HapMap3 variants.^44,45^ Considering the relatively small sample size for other ancestry groups (AFR, AMR, CSA, EAS, and MID), the mixed-linear-model association (MLMA) method was used to perform GWAS using the full GRM.^46^ Additionally, sex-specific GWAS were conducted for each ancestry group using fastGWA for EUR participants and MLMA for other ancestry groups, excluding sex as a covariate from all models.

To explore the influences of potential covariates, we performed GWAS in the EUR ancestry participants using three models. In model 1, we included age, sex, age × sex, and the top 10 genetic principal components derived from the Pan-UKB as covariates. Model 2 added additional covariates, including body mass index, Townsend deprivation index, smoking status, alcohol status, physical activity, and statin use.^47,48^ In model 3, we adjusted for fish oil supplementation status and lipoprotein lipids including chylomicron, very-low-density lipoprotein (VLDL), intermediate-density lipoprotein (IDL), low-density lipoprotein (LDL), high-density lipoprotein (HDL) cholesterols as additional covariates. For GWAS in the non-EUR population, we applied models 1 and 2.

### Identification of novel loci

To identify novel loci, we compared our independent loci to the previously reported FA loci in the GWAS Catalog and in other relevant publications identified by literature review up until June 2024.^49^ Independent loci were identified in our results or previous GWAS based on genome-wide significance threshold (*P*-value < 5 × 10^−8^) and LD-based clumping (*r*^2^ = 0.1, window size = 250 kb) using PLINK.^50,51^ Independent loci were merged based on proximity (±250 kb). We annotated the lead variants for their target genes and functional consequences using the Ensemble GRCh37 Variant Effect Predictor (VEP).^52^

### Heritability and genetic correlation analyses

Heritability and genetic correlations were estimated with linkage disequilibrium score regression (LDSC) in the EUR ancestry subset.^53,54^ In-sample LD scores were derived from s3://broad-alkesgroup-ukbb-ld/UKBB_LD/.^55^ As recommended, we checked for inflation using the attenuation ratio in the EUR population and genomic inflation factor (λ_GC_) in other populations.^56^

### Colocalization with FA traits

We performed colocalization analysis with HyPrColoc (v.1.0) to assess whether two or more FA traits share a putative causal variant.^57^ Pairwise and multi-trait colocalization analyses were conducted for each pair or all FA traits altogether. We used the default priors, including the probability of a variant being associated with a single trait (prior.1 = 1 × 10^−4^) and the conditional probability of association with an additional trait (prior.c = 0.02). A posterior probability (PP) > 0.8 was considered as evidence for a shared causal variant.

### Statistical fine-mapping of GWAS loci

To statistically fine-map the candidate causal variants for genome-wide significant loci associated with each FA trait, we used SuSiE (v.0.12.27).^58^ The analysis was performed for the 1 Mb region surrounding the lead variant of each genetic locus. We utilized in-sample LD information to estimate the correlations among SNPs. The 95% credible set identified potential causal variants based on their PP. Only variants with a PP > 0.8 were reported, with higher PP values indicating more likely to be causal variants.

### Gene-based and gene-set analyses

MAGMA v.1.08, implemented in FUMA, was used to perform gene-based and gene-set analyses.^59,60^ In the gene-based analysis, a total of 19,665 protein-coding genes was tested, and Bonferroni correction was applied to establish the significance threshold (*P* < 0.05/19,665 = 2.54 × 10^−6^). We determined the enrichment of candidate FAs-associated genes in specific biological pathways, cellular components, or molecular functions. A total of 15,481 gene sets (5,497 curated gene sets and 9,984 gene ontology terms) obtained from MsigDB were tested, with the Bonferroni-corrected significance threshold set at *P* < 3.23 × 10^−6^.^61^

### Omics PlEiotRopic Association (OPERA) analysis

To provide mechanistic interpretations of FAs-associated variants, we performed an integrative analysis of six types of multi-omics quantitative trait loci (QTL) summary statistics with GWAS signals using the Bayesian method, Omics PlEiotRopic Association (OPERA).^62^ First, we obtained available molecular quantitative trait loci (xQTL) data, including gene expression QTL (eQTL), protein QTL (pQTL), DNA methylation QTL (mQTL), splicing QTL (sQTL), histone modification QTL (hQTL), and chromatin accessibility QTL (caQTL). We downloaded blood *cis*-eQTLs summary statistics generated from the eQTLGen study (N = 31,684).^63^ The plasma pQTL summary statistics were retrieved from the UKB Pharma Proteomics Project (UKB-PPP) using UKB participants (N = 54,219).^64^ The peripheral blood mQTL data were generated by McRae *et al.* (N = 1,980).^65,66^ We derived summary-level sQTL data from whole blood samples in the Genotype Tissue Expression (GTEx) Project v.8 (N = 670).^67^ The H3K27ac and H3K4me1 hQTL in monocytes were generated from the BLUEPRINT project (N = 200).^68^ The caQTL data in lymphoblastoid cell line samples were estimated by Kumasaka *et al.* (N = 100).^69^ We included the molecular phenotypes with at least one xQTL with *P* < 5 × 10^−8^ and excluded the major histocompatibility complex (MHC) region due to its structural complexity. After filtering molecular phenotypes, we retained 15,785 genes, 2,190 proteins, 94,338 DNA methylation sites, 6,639 RNA splicing events, 18,152 histone marks, and 13,873 chromatin accessibility peaks. The posterior probability of association (PPA) threshold of 0.9 and the multi-exposure heterogeneity in dependent instruments (HEIDI) test threshold of 0.01 were used to determine if a GWAS signal overlaps with the QTL signals for one or multiple molecular phenotypes.

## Results

### Sample characteristics

The primary GWAS comprised up to 239,268 individuals of EUR ancestry. GWAS were also conducted on five additional ancestry groups, including up to 3,352 AFR, 508 AMR, 4,663 CSA, 1,445 EAS, and 865 MID individuals. Among EUR participants, the average age was 57 years and females were more likely to have higher PUFA levels (Table S1). The percentage of female participants in EUR, AFR, AMR, CSA, EAS, and MID populations was 53.9%, 59.7%, 66.9%, 44.8%, 64.6%, and 41.8%, respectively. Approximately 31.6% of individuals reported regular use of fish oil supplements and the ratio of omega-3/omega-6 was around 1:8.

### Identification of genome-wide significant loci for circulating FA traits

We first performed EUR ancestry GWAS for each FA trait using three models, respectively. Overall, we identified between 37 and 124 loci for 19 FA traits in model 2, and between 26 and 48 loci in model 3 (Table S2). Approximately 13% of the loci (ranging from 0– 22%) identified in model 2 were not significant in model 1 (Table S3). However, on average, 62% of the loci (range: 19–79%) from model 2 were not significant in model 3, and 31% (range: 18– 43%) of the loci from model 3 were not identified in model 2 (Table S3). Figure 1A shows an example of the Manhattan plot of GWAS for the absolute concentration of total PUFAs in model 2. Of the 122 significant loci identified in model 2, 30 for total PUFAs were also significant in model 3 and a total of 92 loci identified in model 2 were no longer significant after adjusting for fish oil intake and lipoprotein lipids in model 3 (Figure 1B). We observed similar effect estimates between models 1 and 2, whereas model 3 yielded systematically smaller effect estimates than those from the other two models (Figure S1). To further pinpoint the covariates driving the distinction between models 3 and 2, we conducted GWAS based on model 2 by additionally adjusting for fish oil supplementation in model 2.1 and for lipoprotein lipids in model 2.2. The differences in GWAS results between model 3 and the others were attributed to the additional adjustment for lipoprotein lipids (Table S2). For example, variants in the *LDLR* and *MTTP* genes, known for their strong correlations with lipoprotein lipids,^70,71^ were significant in model 2 but not in model 3 for total PUFAs (Figure 1).

**Figure 1.**
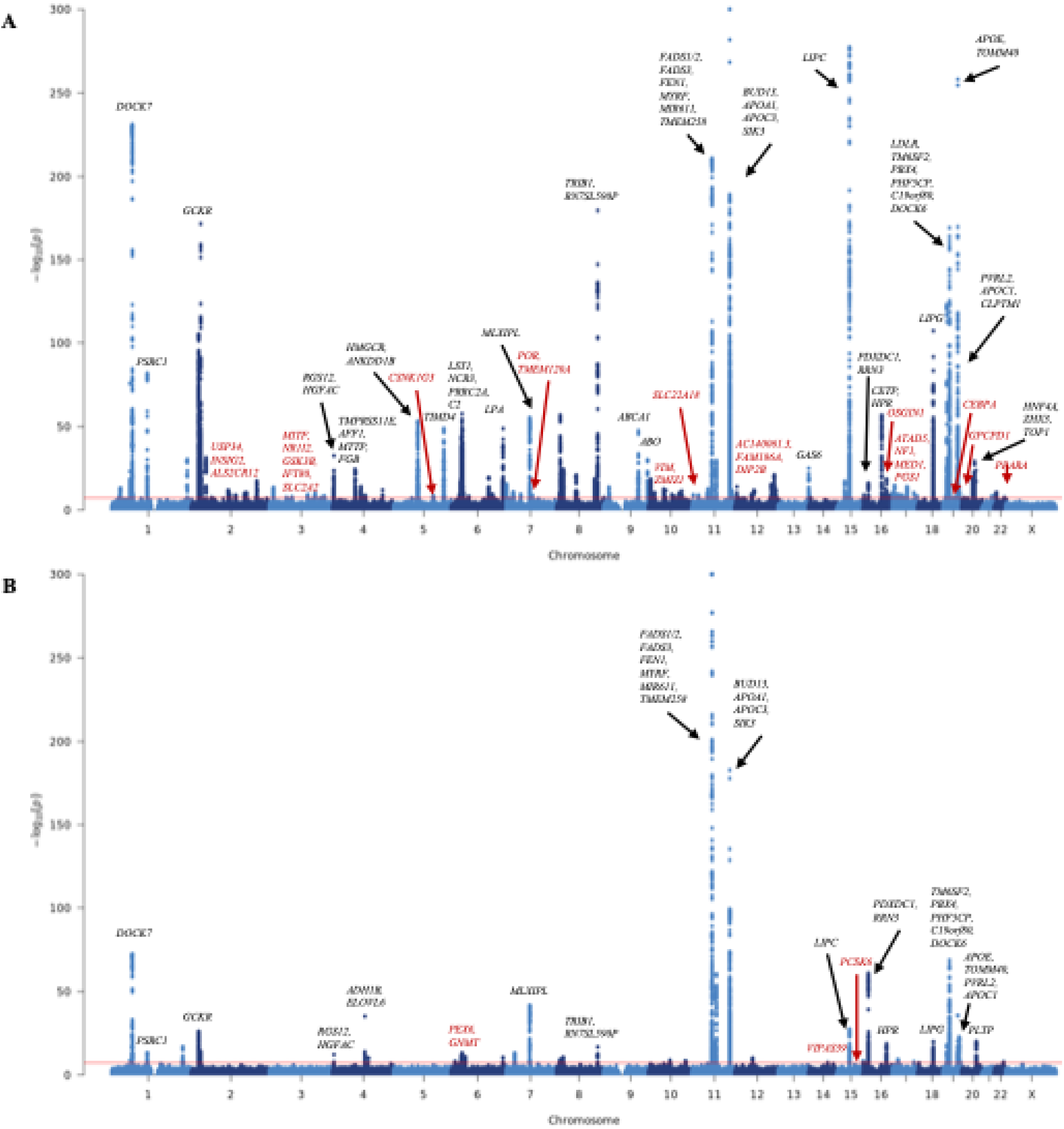
Manhattan plot of the genome-wide association studies for the absolute concentration of total polyunsaturated fatty acids. (A and B) The association of each variant with polyunsaturated fatty acids was obtained from genome-wide association studies using models 2 (A) and 3 (B). A variant with a *P*-value below 5 × 10^−8^ was considered statistically significant. The nearest gene at each genome-wide significant locus is annotated, in black for known loci and in red for novel loci.

Regarding SNP-based heritability of FA traits in the EUR population, we found that SNPs explained 5–19% of the phenotypic variance for all FA traits across all three models (Figure S2A; Table S4). For instance, the SNP-based heritability estimates for the absolute concentrations of total PUFAs, MUFAs, and SFAs were 19%, 19%, and 16%, respectively, in model 2. The genomic inflation factor (λ_GC_) for all GWAS ranged from 1.07 to 1.29 (Table S5). The attenuation ratio ranged from 0.03 to 0.18, and the LDSC intercept ranged from 1.01 to 1.12, suggesting that the genome-wide elevation of association statistics was primarily due to true additive polygenic effects rather than confounders such as population stratification.

To identify novel loci, we searched the literature and GWAS Catalog to compile known loci for FAs-related traits, making it the most comprehensive collection to date (Table S6). When categorizing FA traits into PUFAs-related, MUFAs-related, and SFAs-related, we identified 70, 61, and 54 novel loci among 215, 163, and 119 significant loci, respectively, with model 2 (Table S7). The numbers of novel loci for each FA trait are provided in Table S3. Compared with all previously reported FAs-related loci, the top 10 strongest novel loci identified across all FA traits in model 2 were mapped to candidate genes including *PEPD*, *SBNO1*, *IL1RN*, *INSIG2*, *VIM*, *PGS1*, *ARID5B*, *NF1*, *FAM96A*, and *PEX6* (Table S8). We also compared independent loci to those previously reported in GWAS for the same traits to identify loci that have not been previously associated with the corresponding trait. For example, a novel locus around *VEGFA* was identified for total FAs, LA%, total SFAs, PUFAs/SFAs, and the degree of unsaturation in our study, while it was consistently associated with total MUFAs, MUFAs%, and PUFAs/MUFAs in both our and previous GWAS.^30,33,34,38^ Variants in *GCKR* were found to be significantly associated with DHA% for the first time, while they had been associated with other FA traits (Table S8).

Additionally, we explored genetic associations on the X-chromosome in both overall and sex-specific GWAS. Previously, only one GWAS identified FA loci on the X-chromosome.^30^ That GWAS reported two X-chromosomal loci associated with FA traits; however, no significant variants were identified in our overall GWAS. When evaluating males separately, one significant association with omega-3% was found on the X-chromosome.

### Sex-specific FA GWAS

We then stratified the total sample by sex to perform sex-specific GWAS separately for 128,922 females and 110,346 males in the EUR population. Approximately 4.2% of the significant loci from sex-specific GWAS were not identified in the overall GWAS (Table S3). The SNP-based heritability ranged from 5% to 21% in females and 4% to 20% in males, and there was no evidence of genomic inflation (Figures S2B and S2C; Table S4). In model 2, we identified 128 independent loci associated with PUFAs-related traits in females and 106 loci in males, including 27 and 8 novel loci, respectively, which have not been reported in previous GWAS of PUFAs-related traits (Table S7). GWAS results for both known and novel loci associated with each FA trait are provided in Table S9 for females and Table S10 for males. Some novel loci were only identified in one sex but not in the other. For example, in the model 2 GWAS for total PUFAs, there were three novel loci in females but not in males, when compared to previous GWAS of all FA traits. Two of them were mapped to genes *RNU6-1180P* and *KRT18P55*, while the third locus, located on Chromosome 10, was not mapped to any genes (Figure 2). Similarly, in the model 3 GWAS for total PUFAs, there were three novel loci, around genes *UNC5CL*, *RP11-328J6.1*, and *SIPA1L3*, which were identified only in females (Figure S3). No novel loci were identified in male-specific GWAS for total PUFAs in either model 2 or 3. In the sex-specific GWAS, we found that approximately 43% of the genome-wide significant loci identified in models 1 and 2 were not identified in model 3 (Table S3). On the other hand, about 31% of the loci identified in model 3 were not identified in the other two models (Table S3). Notably, one X-chromosomal locus associated with omega-3%, whose lead SNP is rs147828433, was identified from GWAS using model 2 in males. Our study is the first to explore loci in sex-specific GWAS of FA traits and to include the X chromosome.

**Figure 2.**
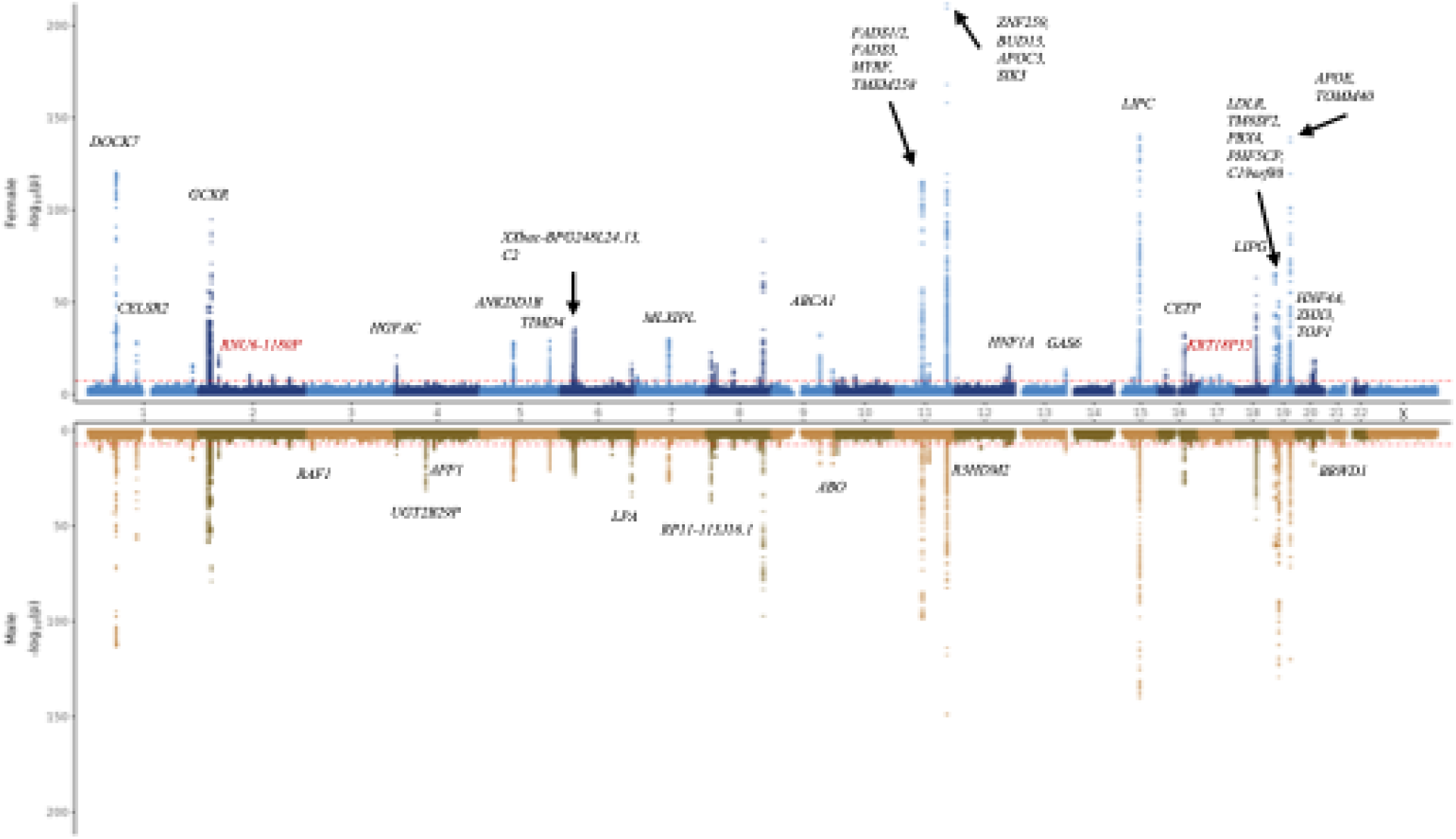
Miami plot of the absolute concentration of total polyunsaturated fatty acids from genome-wide association studies using model 2. The top panel shows the GWAS results in females, while the bottom panel shows the GWAS results in males. The -log_10_(*P*-value) is plotted on the y-axis and chromosomal location is plotted on the x-axis. The genome-wide significance threshold (*P*-value < 5 × 10^−8^) is indicated by the red dashed line. The nearest gene at each genome-wide significant locus is annotated, in black for known loci and in red for novel loci. Note that the lead variant (rs165527) from the novel locus on chromosome 10 (chr10:17259567- 17325281) is not shown in the figure because it is an intergenic variant without an associated gene.

### GWAS for FA traits in individuals of non-EUR ancestries

Across all FA traits, we identified 38 significant loci in ancestry-specific GWAS, of which 23 are novel compared to previous studies in EUR and non-EUR populations (Table S11). The genomic inflation factor (λ_GC_) was also reported for each non-EUR ancestry, with average λ_GC_ values of 0.97, 0.92, 0.99, 0.90, and 0.95 for AFR, AMR, CSA, EAS, and MID, respectively (Tables S5). In the non-EUR ancestry GWAS, the most significant locus was around *FADS2*, identified in four ancestry groups except AFR, which is a well-known locus in the EUR population (Table S11). Among non-EUR populations, CSA had the largest sample size, leading to more loci being identified compared to other groups, with 1–4 loci in model 1 and 0–2 in model 2 (Table S2). The GWAS in CSA male participants identified a novel locus *EDN1* associated with total PUFAs, omega-6 PUFAs, and LA, while this locus did not reach the genome-wide significance in the EUR population. In addition, variants in *VPS39*, which had not been reported in previous GWAS, were associated with total PUFAs, omega-6 PUFAs, and LA in both total and male-only CSA participants.

### Genetic correlations among FA traits

We examined genetic correlations (*r_g_*) between FA traits using EUR GWAS summary statistics from three models (Table S12). Broadly, comparisons of genetic correlations between models revealed highly consistent estimates between models 1 and 2 but more distinct estimates in model 3 (Figure S4). Among the 171 pairs of genetic correlations, 14 pairs that had significantly negative genetic correlations in models 1 and 2 became significantly positive in model 3, while four pairs that were significantly positive in models 1 and 2 became significantly negative in model 3. The strongest correlation was a negative correlation between total MUFAs and PUFAs/MUFAs (*r_g_* = -0.99) observed based on GWAS in both models 1 and 2 (Figures S5 and S6). Notably, genetic correlations between omega-6 PUFAs and omega-6% were negative in models 1 (*r_g_* = -0.38) and 2 (*r_g_* = -0.51), but they were positively correlated in model 3 (*r_g_* = 0.34) after further adjustment for lipoprotein lipids (Figure S7). Within omega-3-related traits, genetic correlations were strong (∼0.9) across all three models, with even stronger correlations found in model 3. We also estimated pairwise phenotypic correlations using Pearson’s correlation coefficient (Table S13). Among significant genetic correlations, all pairs had consistent directions compared to phenotypic correlations in models 1 and 2, while 83% of pairs had consistent directions in model 3, suggesting that genetic effects on FA traits were altered after adding lipoprotein lipids as covariates.

### Colocalization of GWAS signals across FA traits

To assess the probability that two FA traits share the same causal variants, we conducted pairwise colocalization analyses. For the 171 pairs of FA traits, we identified 9–231 colocalized signals in model 2 and 6–114 signals in model 3, respectively. We found that pairs with a larger number of colocalized signals also had stronger genetic correlations (Figure S8). In model 2, potential causal variants in *DOCK7* (rs2934744), *GCKR* (rs1260326), *ZNF259* (rs964184), *FADS1* (rs174564), *CPT1A* (rs2229738), *LIPC* (rs2070895), *LIPG* (rs77960347), *LDLR* (rs73015024), *TM6SF2* (rs58542926), and *APOE* (rs7412) showed robust evidence of colocalization, indicating shared genetic variants for pairs of FA traits (Table S14). After accounting for lipoprotein lipids in model 3, *FADS1* (rs174564), *CPT1A* (rs2229738), and *BUD13* (chr11:116623213:TA:T) were identified as colocalized for FA traits, consistent with results from model 2 (Table S15). Next, we performed multi-trait colocalization analyses to evaluate the posterior probability of a shared genetic signal across all FA traits. Overall, 168 and 159 colocalized signals were identified in models 2 and 3, respectively. We identified *GCKR* (rs1260326), *TRIB1* (rs28601761), *FADS1* (rs174564), *ZNF259* (rs964184), and *APOE* (rs7412) as shared among multiple FA traits in model 2 (Table S16), and *GCKR* (rs1260326), *TRIB1* (rs28601761), *FADS1* (rs174564), and *APOE* (rs1065853) from model 3 as shared signals (Table S17).

### Statistical fine-mapping for candidate causal variants

We applied the Bayesian fine-mapping method to identify putative causal variants for FA traits. We identified 477 unique potential causal variants in model 1, 422 in model 2, and 193 in model 3 (Table S18). Consistent with colocalization analysis, we confirmed that the genetic variants at the loci *TRIB1* (rs28601761), *FADS1* (rs174564), *CPT1A* (rs2229738), *LIPC* (rs2070895), *LIPG* (rs77960347), *LDLR* (rs72658867), *TM6SF2* (rs58542926), and *APOE* (rs7412) are likely to be causally associated with FA traits. Notably, we found that several loci contain multiple 95% credible sets, indicating the existence of multiple potential causal variants within those significant loci. Among the novel loci for FA traits, we found that *ZMIZ1* (rs1782652) was associated with total FAs, total PUFAs, and omega-3 PUFAs, suggesting that it is a candidate shared causal variant for these FAs. Additional potential causal variants for novel loci were identified, including *CPS1* (rs1047891), *ATXN7L1* (rs118061830), *ARID5B* (rs77044968), *SBF2* (rs12789941), *DGKZ* (rs149903077), *CYFIP1* (rs199854211), *FAM96A* (rs62023393), *MIR122* (rs41292412), *BCL2* (rs12454712), *INSR* (rs112630404), and *PEPD* (rs62102718).

### Gene-based and gene-set enrichment analyses

In the genome-wide gene-based association study, 139–541 genes from model 2 and 110–255 genes from model 3 were found to be significantly associated with 19 FA traits after Bonferroni correction (Table S19). The top gene across 19 FA traits identified in model 2 was *ALDH1A2* (chr15:58245622-58790065), which overlaps with another identified gene *LIPC* (chr15:58702768-58861151). In the GWAS of the omega-3/omega-6 ratio using model 3, *STH* was the top gene, which might be related to neurodegenerative diseases such as Alzheimer’s disease.^72^ Gene-tissue expression analysis revealed that the liver was the most significantly enriched tissue, followed by whole blood, kidney, small intestine, spleen, nerve, and adipose tissue (Tables S20 and 21). Additionally, gene-set enrichment analyses were performed to investigate potential gene sets and pathways with enriched GWAS signals (Tables S22 and 23). We found that genes associated with total PUFAs tend to be enriched in biological processes related to lipid homeostasis, secondary alcohol metabolism, organic hydroxy compound transport, regulations of plasma lipoprotein particle levels and lipid metabolism, and fatty acid biosynthesis (Figure 3). Fatty acid metabolic and biosynthetic processes were significantly enriched in the gene-set analysis results from GWAS of total PUFAs using model 3 (Figure S9).

**Figure 3.**
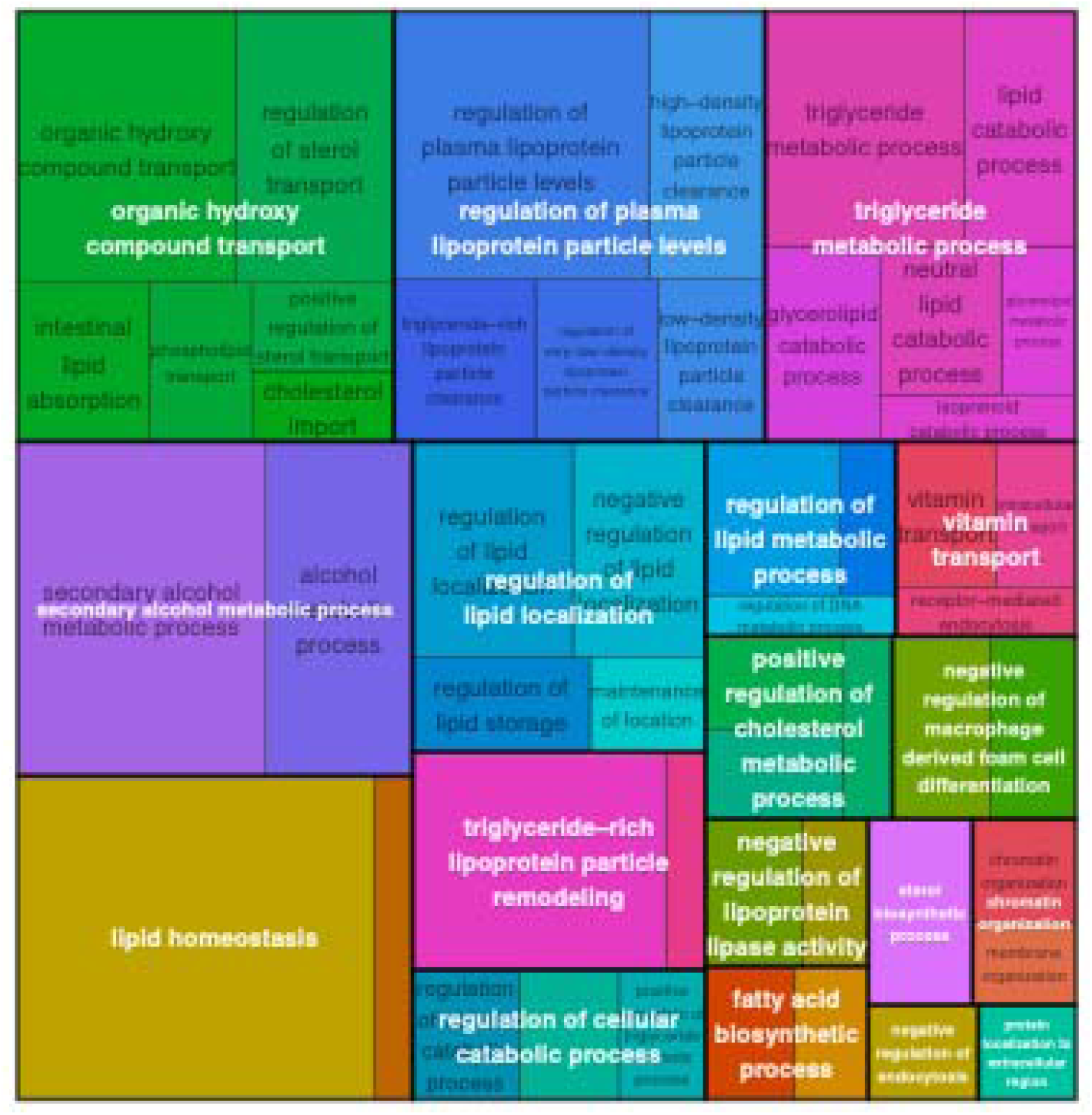
Pathway enrichment analysis of the absolute concentration of total polyunsaturated fatty acids from genome-wide association studies using model 2. Treemap depicting significantly enriched pathways at an adjusted *P*-value threshold of 0.05. Gene ontology terms were clustered based on semantic similarity, with terms displayed as individual rectangles. The color indicates cluster membership, and thick border lines differentiate clusters. The size of each rectangle represents the enrichment significance, and the most significantly enriched term in each cluster is highlighted in white text as the representative term.

### OPERA identifies intermediate molecular phenotypes underlying FAs-associated loci

To better understand the biological mechanisms of our GWAS findings, we applied OPERA to explore the overlap of GWAS signals for FA traits and QTL signals for six molecular phenotypes, including eQTL, pQTL, mQTL, sQTL, hQTL, and caQTL. With a PPA threshold of 0.9 and a *P*_HEIDI_ threshold of 0.01, we identified 976 FAs-associated variants that overlapped with QTL signals for 2,106 unique molecular phenotype measures, including the expression levels of 242 genes, the abundance levels of 60 proteins, the methylation levels at 1,254 DNA sites, the RNA splicing events at 141 sites, the status of 233 histone marks, and the openness at 176 chromatin accessibility peaks (Figure 4A; Table S24). After adjusting for lipoprotein lipids in model 3, we found 481 variants overlapping with xQTLs (Figure 4B; Table S25). Associations with DNA methylation were more frequent than with other molecular phenotypes, and these findings were consistent with previous results in other complex traits.^62^ The number of loci associated with eQTL across 19 traits ranged from 7 for the degree of unsaturation to 68 for total PUFAs in model 2, and from 2 to 41 in model 3. Using the pQTL data from UKB-PPP, we observed that omega-3 PUFAs had the highest number of significant associations with proteins, identifying 35 loci in model 2 and 36 loci in model 3.

**Figure 4.**
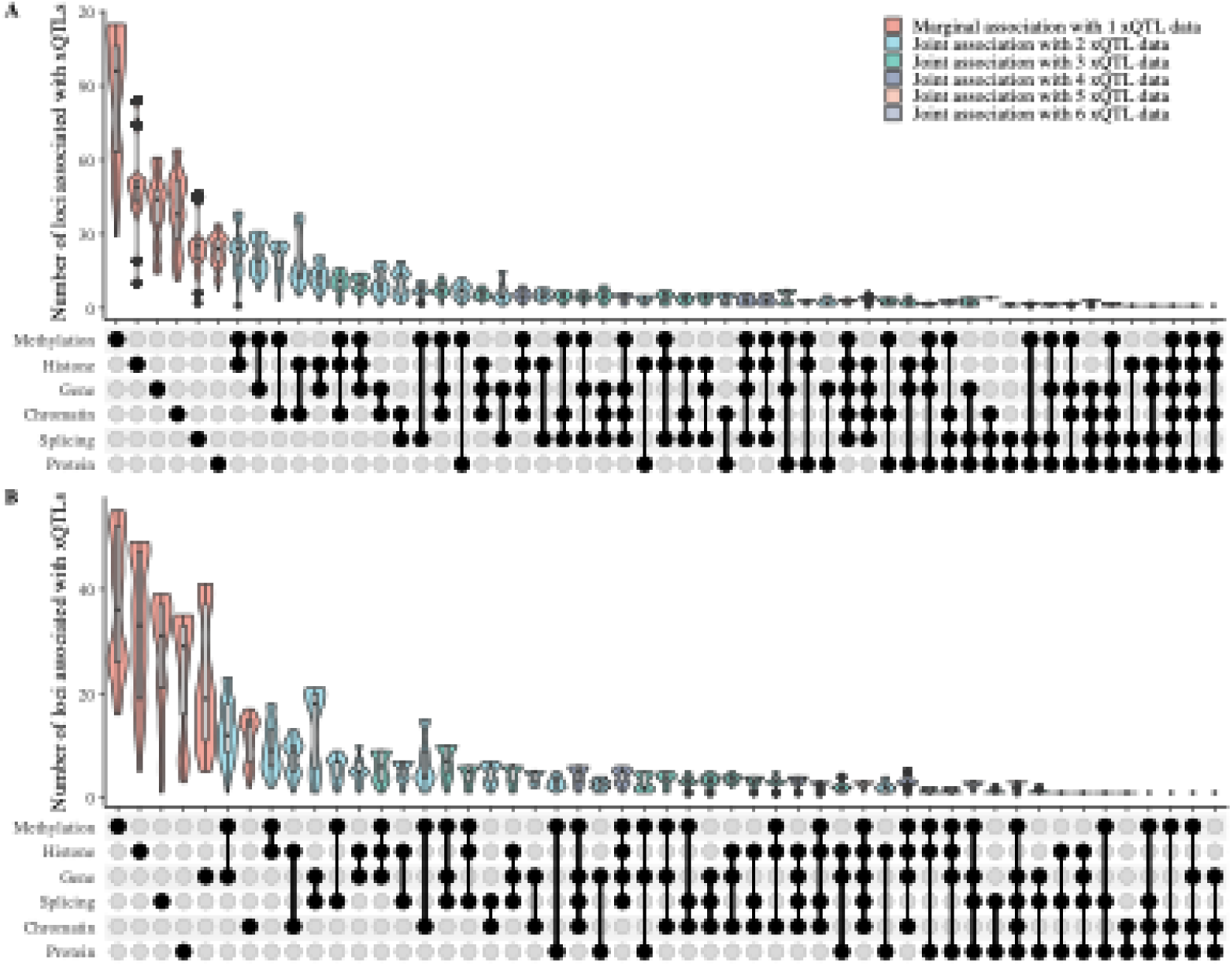
Number of loci for fatty acid traits associated with different molecular phenotype combinations. (A and B) The number of loci associated with 19 fatty acid traits and molecular quantitative trait loci (xQTLs) was determined based on genome-wide association studies using models 2 (A) and 3 (B). The loci numbers are based on significant OPERA association results that pass a posterior probability of association threshold of 0.9 and the multi-exposure heterogeneity in dependent instruments (HEIDI) test (*P*_HEIDI_ > 0.01). The x-axis represents the association hypotheses for different phenotype combinations, while the y-axis shows the number of loci associated with these combinations across 19 fatty acids. Each violin plot displays the distribution of loci numbers by width, with lines indicating the 25^th^ percentile, median, and 75^th^ percentile.

We found that approximately 65% of GWAS loci for 19 FA traits were shared with at least one molecular phenotype. Across FA traits such as total FAs, total PUFAs, omega-6 PUFAs, and total SFAs, the potential causal variant rs72658867, annotated at the *LDLR* gene, was associated with histone modification (chr19:11105519-11214483) and chromatin accessibility (peak 264429). Among the novel loci for omega-6%, rs149903077 (*DGKZ*) was jointly associated with the expression of nearby gene *NR1H3* (ENSG00000025434), histone modification (chr11:47234446-47272168), and chromatin accessibility (peak 200213). It was also marginally associated with methylation (cg01183595).

Notably, all six molecular phenotypes had colocalized QTL signals with omega-6%, total FAs, and total MUFAs at the novel locus *GSTTP1* or nearby genes on chromosome 22 (Figure 5). We observed that the gene *EML3* was associated with the omega-3/omega-6 ratio jointly with five other types of molecular phenotypes, except for histone modification (Figure S10). The nearby variants rs12786457 and rs184864731 were consistently associated with these five phenotypes across GWAS related to omega-3 PUFAs and omega-3%. Overall, OPERA integrates six types of xQTL data to offer mechanistic insights into genetic loci and their downstream molecular phenotypes.

**Figure 5.**
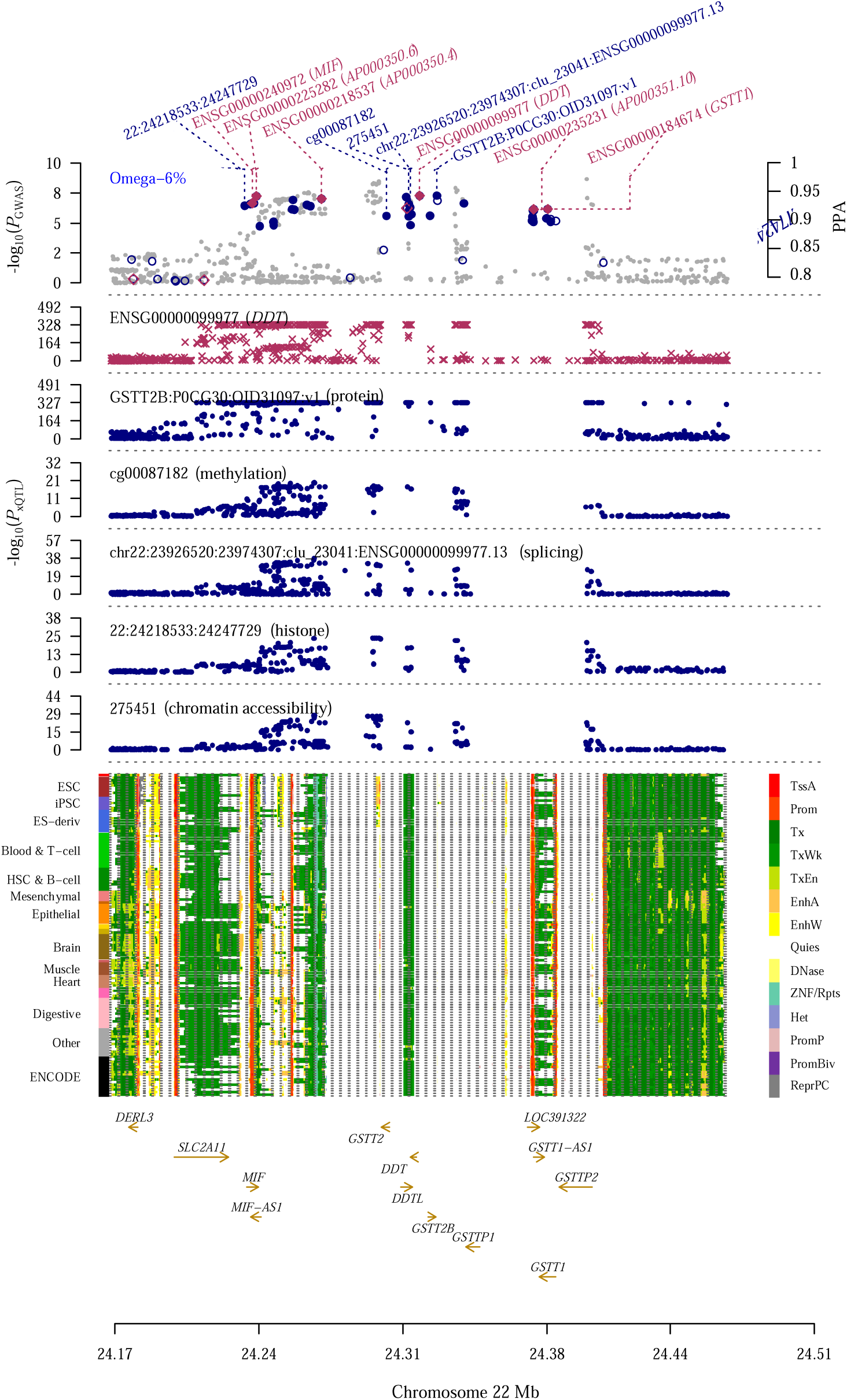
Prioritization of a locus near the *GSTT1/2/2B* genes for the absolute concentration of total fatty acids. The top track displays the -log_10_(*P*-value) of GWAS SNPs (gray dots) for total fatty acids. Red diamonds indicate OPERA marginal PPA for gene associations using eQTL data, while blue circles show OPERA marginal PPA for associations with protein abundance, DNA methylation, RNA splicing, histone modification, and chromatin accessibility, respectively. The bottom track presents 14 chromatin state annotations inferred from the 127 samples of the Roadmap Epigenomics Mapping Consortium.

## Discussion

We performed overall and sex-specific GWAS for 19 FA traits in 250,101 UKB participants of EUR descent and five other ancestry groups, separately. We identified 215, 163, and 119 significant loci for PUFAs-, MUFAs-, and SFAs-related traits, including 70, 61, and 54 novel loci, respectively, in our primary model (i.e., model 2). Further adjustment for lipoprotein lipids in model 3 resulted in significant changes in GWAS signals. On average, 62% of the loci (range: 19–79%) from model 2 were not significant in model 3, while 31% (range: 18–43%) of the loci from model 3 were not identified in model 2. Our analyses of genetic correlations and colocalization revealed the levels of shared genetic basis and specific candidate shared variants across FA traits. Using statistical fine-mapping, we identified 422 putative causal variants across all FA traits. Gene-based and gene-set enrichment analyses pinpointed the liver as the most relevant tissue and highlighted biological pathways underlying FA loci. In addition, we integrated GWAS signals with QTL signals for six molecular phenotypes and revealed that approximately 65% of GWAS loci for 19 FA traits were shared with at least one molecular phenotype, offering novel mechanistic insights.

In EUR GWAS, we analyzed 19 plasma NMR measures of FAs in UKB 239,268 participants. Although previous GWAS of FA traits in the UKB reported many loci, we identified approximately 90% more loci with a doubled sample size.^30,33,34^ In addition, we compared our results with all previous GWAS of FAs-related traits to identify novel loci, including the second-largest published GWAS to date, which included 136,016 participants from 33 cohorts.^38^ We calculated the omega-3/omega-6 and PUFA/SFA ratios for GWAS for the first time. Although the omega-6 to omega-3 ratio is commonly used in studies, the omega-3/omega-6 ratio may better capture the benefits of omega-3. This is particularly relevant given the ongoing debate over the potential harmful or beneficial effects of omega-6 PUFAs.^73^ Our large GWAS of FAs showed that the phenotypic variance explained by all SNPs ranged from 5–19%, while heritability estimates from twin studies were approximately 25–62%.^15^ These results suggest that future studies with rare or structural variants from sequencing and more diverse ancestry groups could be valuable for identifying additional loci for FA traits.

Integrating comprehensive multi-omics data facilitates understanding the mechanisms behind genetic loci and their downstream molecular phenotypes. Our study shows over half of GWAS loci (∼65%) for 19 FA traits are associated with at least one molecular phenotype. For example, the *LDLR* gene variant rs72658867 is linked to histone modification (19:11105519:11214483) and chromatin accessibility (264429), impacting lipid metabolism and cardiovascular health.^74^ The variant rs149903077 in the novel *DGKZ* locus has been associated with the expression of nearby gene *NR1H3*, chromatin accessibility, histone modifications, and DNA methylation, highlighting their combined roles in nuclear receptor regulation, lipid metabolism, and immune responses.^75^ The novel *GSTTP1* locus from the glutathione *S*- transferase (GST) family and nearby genes on chromosome 22 were identified for omega-6%, total FAs, and total MUFAs, overlapping with all six molecular phenotypes and have been linked to key biological processes such as detoxification, cancer susceptibility, and cellular responses to oxidative stress.^76^ These genes in the GST family have previously been reported to be associated with prostate cancer risk, and a recent study found evidence of a nonlinear relationship between omega-6% and prostate cancer.^77–80^ Future studies are needed to validate our findings on the molecular mechanisms underlying FA-relevant variants. These insights could aid in patient stratification for precision nutrition and in identifying novel therapeutic targets related to FAs.

Gene-set enrichment analysis findings highlight biological pathways underlying FA loci, implicating lipid homeostasis, transport, and metabolism, and providing genetic support for the current understanding of fatty acid regulation.^81^ Our previous study suggested that lower omega- 6% may reduce alcohol consumption, with the secondary alcohol metabolism pathway potentially explaining this effect on alcohol-related behaviors.^6^ In model 3, which adjusts for lipoprotein lipid levels, fatty acid metabolic and biosynthetic processes were significantly more enriched in the gene-set analysis of GWAS of total PUFAs. The investigation of the disparities between models 2 and 3 suggests that lipoprotein-related biomarkers play a critical role in FA loci. Future studies are needed to thoroughly dissect these roles, and further analyses, such as Mendelian randomization, should properly infer results using genetic variants with or without adjustment for lipoprotein-related biomarkers.^82^

We investigated the sex-specific genetic architecture of FA traits. A novel locus, *KRT18P55*, was identified in the GWAS for females, and previous research indicates that this locus is highly expressed in patients with gastric cancer.^83^ Notably, only one X-chromosomal locus was identified in the male-only GWAS for omega-3%, tagged by the lead variants rs147828433 (closest protein-coding gene: *ACOT9*). A study on *ACOT9* in mice suggests that it regulates both fatty acid and amino acid metabolism in mitochondria.^84^ For the female-only GWAS, the heritability was higher, and larger numbers of loci were identified. Our results highlight the importance of tracking sex differences in genetics and may motivate future studies on gene-sex interactions. We found higher plasma PUFA levels in females, consistent with previous studies, which were influenced by genetic effects, sex hormones, and conversion rates in FA metabolism.^9^ Randomized controlled trials show sex- and race-specific differences in the benefits of fish oil supplementation and omega-3 PUFAs for preventing cardiovascular events and cognitive decline.^85^

By performing ancestry-specific and sex-specific GWAS in non-EUR samples, we identified additional novel loci, suggesting the presence of ancestry-specific genetic loci for FA traits. In male CSA participants, the novel locus *EDN1*, associated with omega-6 PUFA traits, encodes the protein endothelin 1, which has been linked to cardiovascular events and prognosis.^86^ Variants in *VPS39*, associated with omega-6 PUFAs in the CSA population, have been previously identified as important regulators of myoblast differentiation and muscle glucose uptake in patients with type 2 diabetes.^87^ Besides these ancestry-specific signals, we also found that several loci identified from large EUR GWAS were transferable to other ancestry groups, including *FADS2*, *ZNF259*, *MYRF*, *APOA5*, *GCKR*, *APOE*, and other loci. We found significantly fewer loci in the other five non-EUR ancestry GWAS due to small sample sizes. These results suggest that future studies with larger and more diverse ancestry samples are needed to identify more loci and confirm their consistency across populations.

To the best of our knowledge, our study is the most comprehensive GWAS on FA traits in a very large cohort. First, this is the first study to integrate six types of xQTL data with FA GWAS to explore the possible molecular mechanisms of FAs-associated genetic variants. Second, we used a large number of participants of EUR ancestry with plasma NMR measures for 19 FA traits and included participants from five other ancestry groups. Third, the inclusion of the omega-3/omega-6 ratio in the GWAS provides better insights into the benefits of omega-3 PUFAs. Fourth, our sex-specific GWAS on EUR and non-EUR populations, as well as the inclusion of X-chromosomal, uncover sex differences in FA genetic basis. Fifth, we comprehensively compared our findings to all previous GWAS of FAs-related traits. Detailed information on all novel and known loci has been provided (Tables S3 and S6), which offers a valuable resource for the FA research community. Finally, we adjusted for various covariates across three models, with a particular focus on genetic architecture adjusted for lipoprotein metabolism.

The study has limitations. The numbers of participants in non-EUR populations were small, and future studies with larger sample sizes are necessary. While we explored the molecular mechanisms of FA loci, we are eager to extend OPERA analysis to other tissue-specific molecular phenotypes, particularly the liver, and even specific cell types, to identify cell type-specific disease mechanisms.

In conclusion, our findings reveal novel genetic loci and provide the first evidence of molecular mechanisms underlying FAs-relevant variants. These insights could aid population stratification for precision nutrition and the identification of novel therapeutic targets for FAs-related diseases.

### Data availability

The full summary statistics for GWAS of fatty acids are publicly available in the GWAS Catalog database (https://www.ebi.ac.uk/gwas/, GCP ID: TBA after peer review). The blood eQTL data of eQTLGen Consortium used in the analyses were downloaded from https://molgenis26.gcc.rug.nl/downloads/eqtlgen/cis-eqtl/SMR_formatted/cis-eQTL https://zenodo.org/records/7951839/files/LBC_BSGS_meta.tar.gz?download=1SMR_20191212.tar.gz. UKBB pQTL data are available at https://metabolomips.org/ukbbpgwas/. The mQTL data are available for download from https://zenodo.org/records/7951839/files/LBC_BSGS_meta.tar.gz?download=1. Blood sQTL data can be downloaded at https://yanglab.westlake.edu.cn/data/SMR/GTEx_V8_cis_sqtl_summary/sQTL_Whole_Blood.zip. The hQTL data can be assessed from http://ftp.ebi.ac.uk/pub/databases/blueprint/blueprint_Epivar/qtl_as/QTL_RESULTS/. Summary statistics of caQTL can be found at https://zenodo.org/records/1405945/files/.

## Supplemental information

Supplemental information can be found online at https://doi.org/.

## Acknowledgments

This research was approved by the University of Georgia Institutional Review Board and the UK Biobank consortium (application no. 48818). This work was funded by the University of Georgia Research Foundation and by the National Institute of General Medical Sciences of the National Institutes of Health under Award Number R35GM143060. The content is solely the responsibility of the authors and does not necessarily represent the official views of the National Institutes of Health.

## Author contributions

Y.S. and K.Y. designed the study. Y.S. and H.X. performed analyses. Y.S. and K.Y. drafted the manuscript. All authors contributed to the review and critical revision of the manuscript.

