## Supplementary Figures for "Genome-wide association studies and multi-omics integrative analysis reveal novel loci and their molecular mechanisms for circulating polyunsaturated, monounsaturated, and saturated fatty acids"

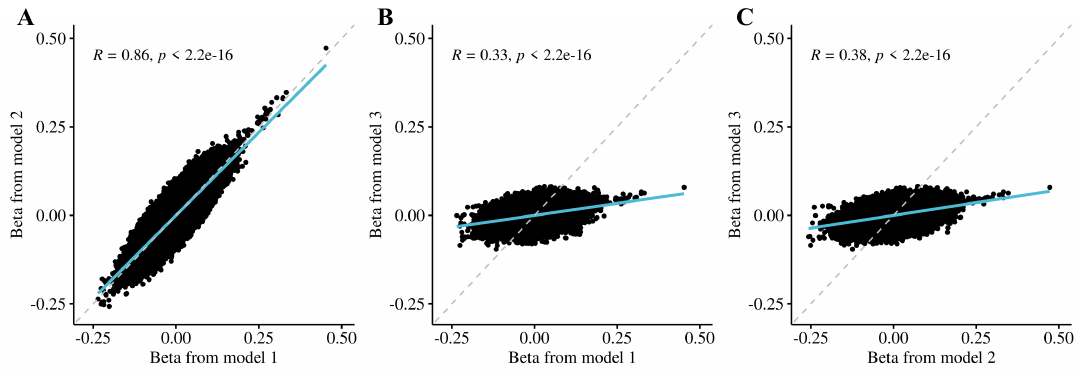
**Figure S1: Correlation between the effect sizes estimated in the genome-wide association studies for the absolute concentration of total polyunsaturated fatty acids between models.**

Pearson correlation (*R*) of effect sizes of SNPs in GWAS between models 1 and 2 (A), 1 and 3 (B), and 2 and 3 (C), respectively.

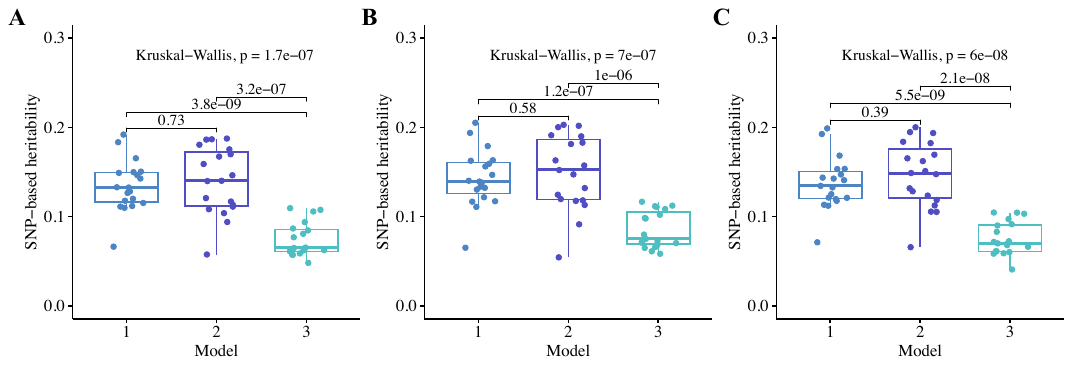
**Figure S2: Boxplots of SNP-based heritability estimates for all fatty acid traits using three models.**

The SNP-based heritability estimates are shown for the overall GWAS (A), sex-specific GWAS for females (B), and sex-specific GWAS for males (C).

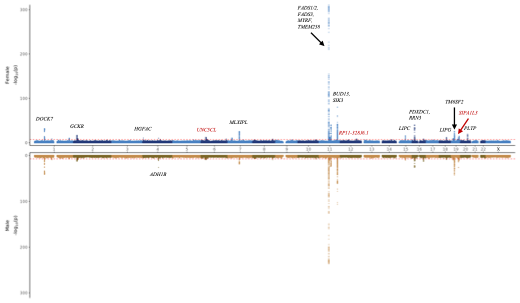
**Figure S3: Miami plot of the absolute concentration of total polyunsaturated fatty acids from genome-wide association studies using model 3.**

The top panel shows the GWAS results in females, while the bottom panel shows the GWAS results in males. The -log_10_(*P*-value) is plotted on the y-axis and chromosomal location is plotted on the x-axis. The genome-wide significance threshold (*P*-value < 5 × 10^−8^) is indicated by the red dashed line.

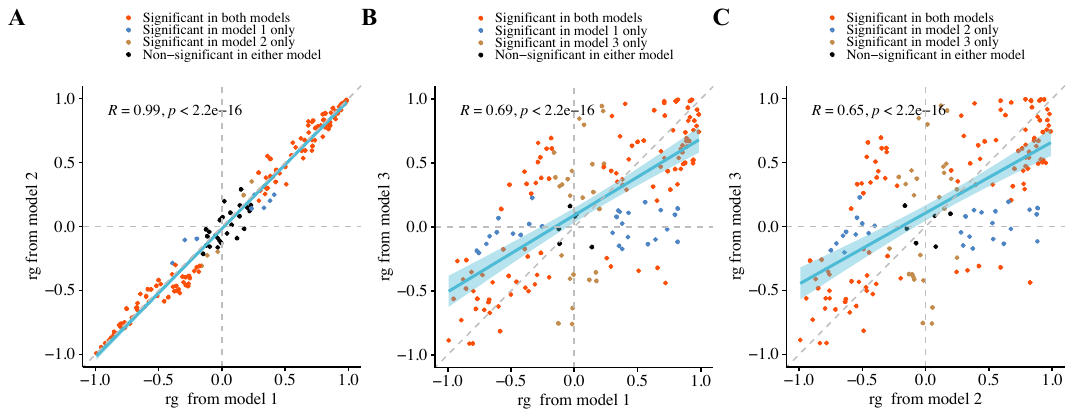
**Figure S4: Scatter plots of genetic correlations across 19 fatty acid traits comparing three models.**

Genetic correlations across 19 fatty acid GWAS between models 1 and 2 (A), 1 and 3 (B), and 2 and 3 (C), respectively.

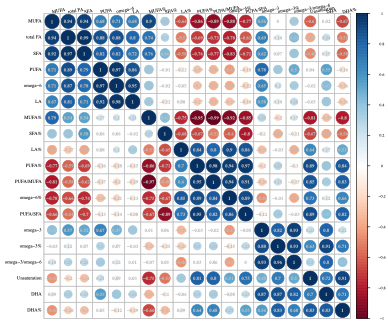

**Figure S5: Genetic and phenotypic correlations among fatty acid traits based on genome-wide association studies from model 1.**

The upper and lower triangles display pairwise genetic and phenotypic correlation estimates. PUFAs, polyunsaturated fatty acids; DHA, docosahexaenoic acid; LA, linoleic acid; omega-3/omega-6, ratio of omega-3 to omega-6 PUFAs; FAs, fatty acids; MUFAs, monounsaturated fatty acids; SFAs, saturated fatty acids. The "%" represents the relative percentage of total fatty acids.

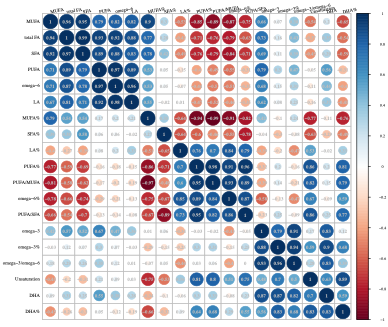

**Figure S6: Genetic and phenotypic correlations among fatty acid traits based on genome-wide association studies from model 2.**

The upper and lower triangles display pairwise genetic and phenotypic correlation estimates. PUFAs, polyunsaturated fatty acids; DHA, docosahexaenoic acid; LA, linoleic acid; omega-3/omega-6, ratio of omega-3 to omega-6 PUFAs; FAs, fatty acids; MUFAs, monounsaturated fatty acids; SFAs, saturated fatty acids. The "%" represents the relative percentage of total fatty acids.

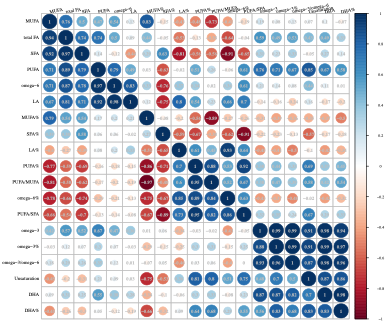

**Figure S7: Genetic and phenotypic correlations among fatty acid traits based on genome-wide association studies from model 3.**

The upper and lower triangles display pairwise genetic and phenotypic correlation estimates. PUFAs, polyunsaturated fatty acids; DHA, docosahexaenoic acid; LA, linoleic acid; omega-3/omega-6, ratio of omega-3 to omega-6 PUFAs; FAs, fatty acids; MUFAs, monounsaturated fatty acids; SFAs, saturated fatty acids. The "%" represents the relative percentage of total fatty acids.

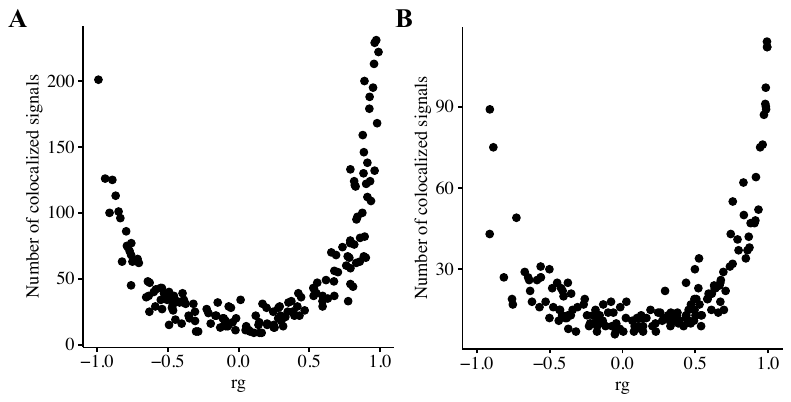

**Figure S8: Scatter plots of genetic correlations and pairwise colocalization analyses across 19 fatty acid traits.**

Genetic correlations and pairwise colocalization analyses across 19 fatty acid GWAS using models 2 (A) and 3 (B), respectively.

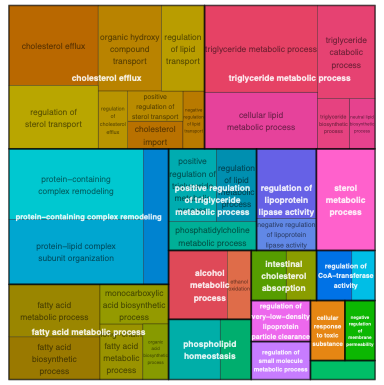

**Figure S9: Pathway analysis of the absolute concentration of total polyunsaturated fatty acids from genome-wide association studies using model 3.**

Treemap depicting significantly enriched pathways at an adjusted *P*-value threshold of 0.05. Gene Ontology terms were clustered based on semantic similarity, with terms displayed as individual rectangles. The color indicates cluster membership, and thick border lines differentiate clusters. The size of each rectangle represents the enrichment significance, and the most significantly enriched term in each cluster is highlighted in white text as the representative term.

**Figure S10: Prioritization of a locus near the *EML3* gene for the ratio of omega-3 to omega-6 PUFAs.**

The top track displays the -log_10_(*P*-value) of GWAS SNPs (gray dots) for total fatty acids. Red diamonds indicate OPERA marginal PPA for gene associations using eQTL data, while blue circles show OPERA marginal PPA for associations with protein abundance, DNA methylation, RNA splicing, and chromatin accessibility, respectively. The bottom track presents 14 chromatin state annotations inferred from the 127 samples of the Roadmap Epigenomics Mapping Consortium.
